# Genome-wide association study of blood mercury in European pregnant women and children

**DOI:** 10.1101/2023.02.06.23285518

**Authors:** Kyle Dack, Mariona Bustamante, Caroline M. Taylor, Sabrina Llop, Manuel Lozano, Paul D Yousefi, Regina Grazuleviciene, Kristine Bjerve Gutzkow, Anne Lise Brantsæter, Dan Mason, Georgia Escaramís, Sarah J Lewis

**Affiliations:** Medical Research Council Integrative Epidemiology Unit, University of Bristol, Bristol BS8 1TH, UK; ISGlobal, Institute for Global Health, Barcelona, Spain; Universitat Pompeu Fabra (UPF), Barcelona, Spain; Spanish Consortium for Research on Epidemiology and Public Health (CIBERESP), Madrid, Spain; Centre for Academic Child Health, Bristol Medical School, University of Bristol, Bristol BS8 2PS, UK; Epidemiology and Environmental Health Joint Research Unit, FISABIO-Universitat Jaume I- Universitat de València, Valencia, Spain; Preventive Medicine and Public Health, Food Sciences, Toxicology and Forensic Medicine Department, Universitat de València, Valencia, Spain; Department of Environmental Sciences, Faculty of Natural Sciences, Vytautas Magnus University, 53361 Academia, Lithuania; Department of Air quality and noise, Division of Climate and Environmental Health, Norwegian Institute of Public Health, P.O. Box 222 Skoyen, NO-0213 Oslo, Norway; Department of Food Safety, Division of Climate and Environmental Health, Norwegian Institute of Public Health, P.O. Box 222 Skoyen, NO-0213 Oslo, Norway; Bradford Teaching Hospitals NHS Foundation Trust, Duckworth Lane, Bradford BD9 6RJ, UK; CIBER in Epidemiology and Public Health (CIBERESP), Barcelona, Spain; Department of Biomedical Sciences, Institute of Neuroscience, University of Barcelona, Barcelona, Spain; Population Health Sciences, Bristol Medical School, University of Bristol, Bristol BS8 1TH, UK

**Keywords:** GWAS, blood mercury, pregnant women, children, ALSPAC, HELIX

## Abstract

**Background:** Mercury (Hg) is a toxic heavy metal which humans are most commonly exposed to through food chain contamination, especially via fish consumption. Even low-level exposure can be harmful because of the poor clearance rate, particularly for methylmercury. It is likely that genetic variation modifies exposure through changes in the absorption, metabolism, and/or removal of mercury. Associations have been reported between Hg and variants at multiple genetic loci, but in many cases these results are not yet replicated.

**Methods:** This study included two populations: pregnant women from the Avon Longitudinal Study of Parents and Children (ALSPAC, n=2,893) and children from the Human Early Life Exposome (HELIX, n=1,042). Genome-wide testing by cohort was performed by fitting linear regressions models on whole blood Hg levels and Haplotype Reference Consortium imputed single-nucleotide polymorphisms (SNPs). SNP heritability was estimated using linkage disequilibrium (LD)-score regression, and the biological functions of the top variants were investigated using resources which aggregate prior literature.

**Results:** Hg SNP heritability was estimated to be 24.0% (95% CI: 16.9% to 46.4%) for pregnant women. The number of genetic variants independently associated with whole blood mercury levels above a suggestive p-value threshold (P < 1×10^−5^) was 16 for pregnant women and 21 for children. However, none were replicated in both populations, nor did any pass a stronger genome-wide significant threshold (P < 5×10^−8^). Several suggestive variants had possible biological links to Hg such as rs146099921 in metal transporter *SLC39A14*, and two variants (rs28618224, rs7154700) in potassium voltage-gated channels genes.

**Discussion:** There was evidence for a considerable proportion of Hg variance being attributed to genome-wide variation in pregnant women. However, results between pregnant women and children were highly discordant which could reflect differences in metabolism and a gene-age interaction with Hg levels. There were a large number of SNPs suggestively associated with Hg levels, which likely include both true associations and false positives. These interim findings will be expanded following collaboration with additional study groups.

## Introduction

Mercury (Hg) is a toxic metal which has increased in environmental concentrations substantially over the past century, the increase largely being attributed to changing human activities (1). Hg can be found in three forms, elemental, inorganic (I-Hg), and organic mercury mainly in the form of methylmercury (MeHg) (2). Of these, MeHg is considered to be the most harmful to humans, especially in vulnerable populations such as young children (3).

I-Hg is released in atmospheric emissions from industrial processes such as fossil fuel burning and transported globally through the atmosphere and oceans (4, 5). MeHg is formed as a result of the biomethylation of I-Hg due to the action of bacteria present in the aquatic environment (6). The amount of MeHg found in fish increases along the food chain because of bioaccumulation and biomagnification, with greater levels being observed in species occupying higher trophic levels, as well as in older specimens. While Hg primarily enters the human food chain through seafood contamination (7, 8), it can also contaminate rice, cereals (9, 10), meat (11), and be found in small quantities in other food products (12).

The properties of mercury exposure and metabolism are tissue and compound dependent. Circulating blood Hg is primarily composed of MeHg, with the remainder being I-Hg (13), and commonly the two are measured together as total Hg. MeHg is primarily absorbed through diet (14), and blood Hg can therefore be indicative of recent dietary exposure. Absorbed I-Hg tends to bind to thiol-containing proteins, and is quickly transported by plasma proteins such as albumin out of circulation and into tissues throughout the body (15). MeHg on the other hand is largely circulated in erythrocytes, potentially by binding to the erythrocyte membrane, or through other transport mechanisms such as D-glucose or cysteine (15, 16). A more detailed description is discussed elsewhere (15). The majority of MeHg excretion follows its demethylation to I-Hg (17), after which it may be transported to the kidneys or secreted from blood into the intestines for excretion through urine or faeces respectively (18, 19).

Mercury once absorbed is structurally similar to common amino acids (20), which enables binding with sulfhydryl group organic compounds, lipids, proteins, and enzymes. The toxic effects of Hg are therefore broad and can harm cellular functioning and health in any tissue (21). Macro-biological consequences include increased reactive oxygen species and oxidative stress (22, 23), carcinogenesis (24), epigenetic changes (25), and cell death (26), among many others.

The toxicity of Hg means that high doses of exposure such as from occupational accidentals can lead to acute poisoning, and require immediate treatment to avoid organ failure, neurological impairment, and long-term harm (27-29). For the remainder of the population exposure is mainly through diet, which involves smaller doses of Hg but can still cause to long-term harm. This is because Hg is slow to clear from the body and over time toxic quantities may accumulate within the body. Mercury levels are associated with increased blood pressure (30), risk of heart disease (31), and kidney disease (32), and a wide range of neurological symptoms (14, 33). Babies and children are predicted to be especially vulnerable, because mercury can readily cross the placenta (34) and accumulate at a higher Hg-to-weight ratio in the child. However, the evidence linking lower-level mercury exposure to developmental harm is currently weak (35, 36).

The identification of genetic variants associated with mercury concentrations could enable new methods of assessing the health effects of exposure, by acting as randomized proxies of mercury exposure (37). Genetic variation in the form of single-nucleotide polymorphisms (SNPs) could alter the rates of absorption, transport, tissue storage, or removal of mercury. Human genetic association studies have identified several genes where variation could impact Hg, such as genes involved in the glutathione metal-binding detoxification system (38-40), the metallothionein metal transport family (41), lipid-transport protein APOE (42), and iron homeostasis (43). Metal-metal interactions occur with selenium (44), zinc (45, 46), cadmium, and lead (47, 48) and therefore variants linked to these metals may also affect mercury levels (49-51).

Hypothesis-free genome-wide association studies (GWAS) allow systematic assessment of the relationship between single nucleotide polymorphisms (SNP) and phenotypes of interest where results are easy to meta-analyse through standardised summary data and variants can be identified not only in genes but also non-coding regions of the genome (52, 53). This method has previously been used to identify genetic variants associated with blood concentrations of copper (50), iron (54) lead (49), manganese (55), selenium (50, 51), and zinc (50).

The objective of this study was to assess the associations between SNPs and blood Hg concentrations in pregnant women and children in a GWAS. Specifically, we aimed (1) to estimate the heritability of blood Hg levels, (2) to identify genetic variants associated with Hg levels in two European populations using genome-wide association testing and explore their functions through in silico analyses, and (3) to determine associations between Hg levels and candidate variants reported in previous studies of Hg or linked metals.

## Methods

### Overview

Genome-wide associations were estimated between genetic variants and blood Hg concentrations in two separate European studies, one of pregnant women and one of children. An overview of study characteristics, analysis methods, and exclusions is provided in Supplementary Table S1.

### The Avon Longitudinal Study of Parents and Children (ALSPAC)

ALSPAC is a multi-generational birth cohort in the former Avon Health Authority area in the UK. All pregnant women living within this area with expected dates of delivery between 1^st^ April 1991 and 31^st^ December 1992 were invited to take part in the study. From 20,248 pregnancies identified as eligible, 14,541 were initially enrolled which after accounting for multiple pregnancies resulted in 14,203 unique mothers. This was expanded with additional phases of recruitment to provide a total of 14,833 unique women in the study. Full details of the recruitment process and sample profile are described elsewhere (56, 57). Details of all the data that is available from the study are available in a fully searchable online data dictionary and variable search tool: http://www.bristol.ac.uk/alspac/researchers/our-data/. Participant characteristics were representative of the majority of UK women. However, women were predominantly of European ancestry, and only those women were included in this GWAS analysis, which limits the generalizability of findings to populations with different ancestry (56).

Whole blood samples were taken from 4,844 pregnant women during early antenatal care visits, with a median visit time of 11 weeks of gestation (IQR: 4 weeks). A vacutainer system was operated by midwives to draw the samples, which were stored at 4°C for 1-4 days before being sent to the central Bristol laboratory. Samples were transported for up to 3 hours at room temperature, and then stored at 4°C until the time of analysis.

Whole blood Hg was measured using inductively coupled plasma dynamic reaction cell mass spectrometry (ICP-DRC-MS) at the Centers for Disease Control and Prevention (CDC), Bethesda, CDC method 3009.1. Quality control measures are described in earlier studies (50, 58), which after exclusions left 4,131 measurements. One sample was below the limit of detection for Hg (0.24 μg/L) and was assigned a value 0.7 times the lower limit of detection (59).

Blood samples for DNA analysis were taken during pregnancy from 10,015 women (60). Samples were genotyped by Centre National de Génotypage (CNG) using the Illumina Human660W-Quad Array. Genotype annotation was performed using Illumina GenomeStudio (61), and aligned to GRCh37 with the software Burrows-Wheeler Aligner. Quality control procedures were applied to the genotyped data using Plink v1.07 (62). SNPs were excluded if they were missing from more than 5% of individuals, had a Hardy-Weinberg (HWE) equilibrium P < 1.0×10^−07^, or a minor allele frequency (MAF) of less than 1%. Individuals were excluded if they were missing more than 5% of SNPs, had indeterminate X chromosome heterozygocity or extreme autosomal heterozygocity (> 3 standard deviations from population mean), were population outliers using four HapMap populations as reference, or had a cryptic relatedness estimate equivalent to first cousin or closer (IBD > 0.125) with another individual in the sample (63, 64). Directly genotyped SNPs were imputed to the Haplotype Reference Consortium (HRC r1.1) panel of approximately 31,000 phased whole genotypes. Phasing was performed using ShapeIt v2 (65) and imputation using Impute V3 on the Michigan Imputation Server (66). SNPs were excluded following imputation where MAF <1% or imputation quality score (INFO) < 0.9.

### The Human Early-Life Exposome (HELIX)

HELIX comprises subcohorts of mother-child pairs from six European birth cohorts (67, 68). The cohorts enrolled approximately 32,000 pairs between 1999 and 2010 in the UK, France, Spain, Lithuania, Norway (69), and Greece. From these studies, 1,301 children were included in HELIX subcohort which measured a variety of pre- and postnatal exposures, health outcomes, and genome-wide genotypes. The current study only included children with genetic data, Hg levels, and who were of European ancestry (determined from genome-wide genetic information) (n=1,042).

Child blood samples were collected during follow-up clinic visits between December 2013 to February 2016, when the children were aged 6 to 12 years old (70). All cohorts followed the same procedures and analysis protocols. Whole blood was stored in EDTA vacutainers, and analysed for trace element testing and DNA extraction at ALS Scandinavia (Sweden). Total Hg levels were measured using double focusing sector field inductively coupled plasma mass spectrometry (ICP-SFMS) as described elsewhere (71). The limit of detection was 0.02 μg/L.

The Infinium Global Screening Array (GSA) (Illumina) was used for genome-wide genotyping at the Human Genomics Facility (HuGe-F), Erasmus MC (www.glimdna.org). GenomeStudio software with the GenTrain2.0 algorithm was used for genotype calling, and annotation on GRCh37 using the GSAMD-24v1-0_20011747_A4 manifest. Samples were excluded if there was SNP missingness >3%, sex mismatch, heterozygosity (>4 SD), cryptic relatedness (Pi-hat > 0.185), or duplicates. SNPs were excluded if missing from >5% individuals, MAF < 1%, or HWE P < 1.0×10^−06^.

Genotype information was phased to HRC r1.1 using Eagle v2.4, and imputed using Minimac4 and the Michigan Imputation Server (72). Post imputation filtering was applied to exclude SNPs with low imputation quality (R2 < 0.9), allele frequency (MAF < 1%), or HWE P > 1.0×10^−07^.

### Estimation of SNP-heritability

The LD Score regression tool (73, 74) was used to estimate phenotype SNP heritability (h^2^g) according to the summary statistics of each GWAS. In brief, the method involved regressing summary statistics from each GWAS study against an LD score reference panel computed using 1000 Genomes Project European data (75). The reference panel was accessed from https://data.broadinstitute.org/alkesgroup/LDSCORE with the filename ‘eur_w_ld_chr’.

### Genome wide association studies

Genome-wide association testing was performed to estimate the association between each SNP and a continuous Hg phenotype. In ALSPAC, the GWAS of pregnant women was conducted in SNPTEST version 2.5.2 using the frequentist option and “score” method of accounting for genotype uncertainty (76). In HELIX the GWAS of children was ran using PLINK version 1.0 with the “--linear” option (62). Each analysis adjusted for age at the time when blood was taken and eigenvectors for the first 10 principal components estimated from GWAS data. The continuous linear models assumed an additive effect of SNPs and a normal distribution of phenotype residuals. The distribution of Hg had a strong right skew and if regressed in its raw form was unlikely to meet the latter assumption, and this could have biased the model standard error and p-values. To address this, Hg measurements were log2 transformed to approximate a normal distribution.

Follow-up analysis was performed in R version 4.1.0 unless otherwise stated. The reference SNP IDs were missing from all ALSPAC results and some HELIX, and therefore chromosome and location were used to identify ID labels valid for GRCh37 using the “*SNP locations for Homo sapiens (dbSNP Build 144)”* reference table and “*BSgenome”* R packages (77, 78). Results were visualised with quantile-quantile (QQ) and Manhattan plots generated using the “*qqman*” package (79). SNPs were classified as genome-wide significant if p< 5×10^−8^, and suggestively significant if p was between 1×10^−5^ and 5×10^−8^. Variants in linkage disequilibrium (R^2^>0.1 and 250 kb range) were grouped using the ld_clump function of the MRC IEU GWAS R package (80) and the most significant SNP kept. The strongest results from each GWAS were compared.

In addition to identifying suggestive and significant SNPs, summary statistics were extracted and reported for 13 variants of interest which were previously identified as (a) associated with Hg levels in candidate gene studies or (b) associated with metals which may interact with Hg levels in genome-wide association studies (Supplementary Table S2).

### In silico functional analysis

All variants with p < 1×10^−5^ were mapped to the nearest gene using Functional Mapping and Annotation of Genome-Wide Association Studies (FUMA) SNP2Gene function (81) and NCBI Sequence Viewer (82). The potential biological mechanisms of how the variants may affect Hg were investigated using tools that aggregated prior genetic research. Variant-phenotypes associations were explored using FUMA eQTL (81), LDtrait (83) and PhenoScanner (84, 85). Gene functions were explored in GeneCards database (86), tissue expression using GTEx portal (87, 88), and gene-phenotype associations in the Online Mendelian Inheritance in Man database (OMIM) (89).

## Results

### Study characteristics

There were 2,893 pregnant women included in this study, from the wider ALSPAC population with both Hg measurements (total n = 4,014) and imputed genotype (n = 8,196) data. From HELIX there were 1,042 children included, from 1,298 Hg and 1,303 imputed genotype measurements. Full sample size derivation is provided in Supplementary Table S3.

Pregnant women were a median age of 28.0 years (IQR: 6.0), and children 8.0 years (IQR: 2.4). The child population were 54.6% male and from six cohorts and countries (Table 1).

**Table 1.** Composition of the study populations.

|  | Country | Size |
| --- | --- | --- |
| **Children (HELIX)** | - | 1,042 |
| Born in Bradford (BiB) | United Kingdom | 90 |
| Étude des Déterminants pré et postnatals du développement et de la santé de l'ENfant (EDEN) | France | 135 |
| INfancia y Medio Ambiente (INMA) | Spain | 198 |
| Kaunas birth cohort (KANC) | Lithuania | 196 |
| The Norwegian Mother, Father and Child cohort study (MoBa) | Norway | 237 |
| The Rhea Mother-Child Cohort in Crete (Rhea) | Greece | 186 |
| **Pregnant women (ALSPAC)** | United Kingdom | 2,983 |

For pregnant women, the mean Hg was 2.09 μg/L (standard deviation, SD: 1.08) and median 1.89 μg/L (interquartile range, IQR: 1.16). There was a lower mean (1.35 μg/L) and median (0.82 μg/L) for children, but greater variance (IQR: 1.27). As seen in Figure 1, in both studies the distribution of Hg was right skewed.

**Figure 1.**
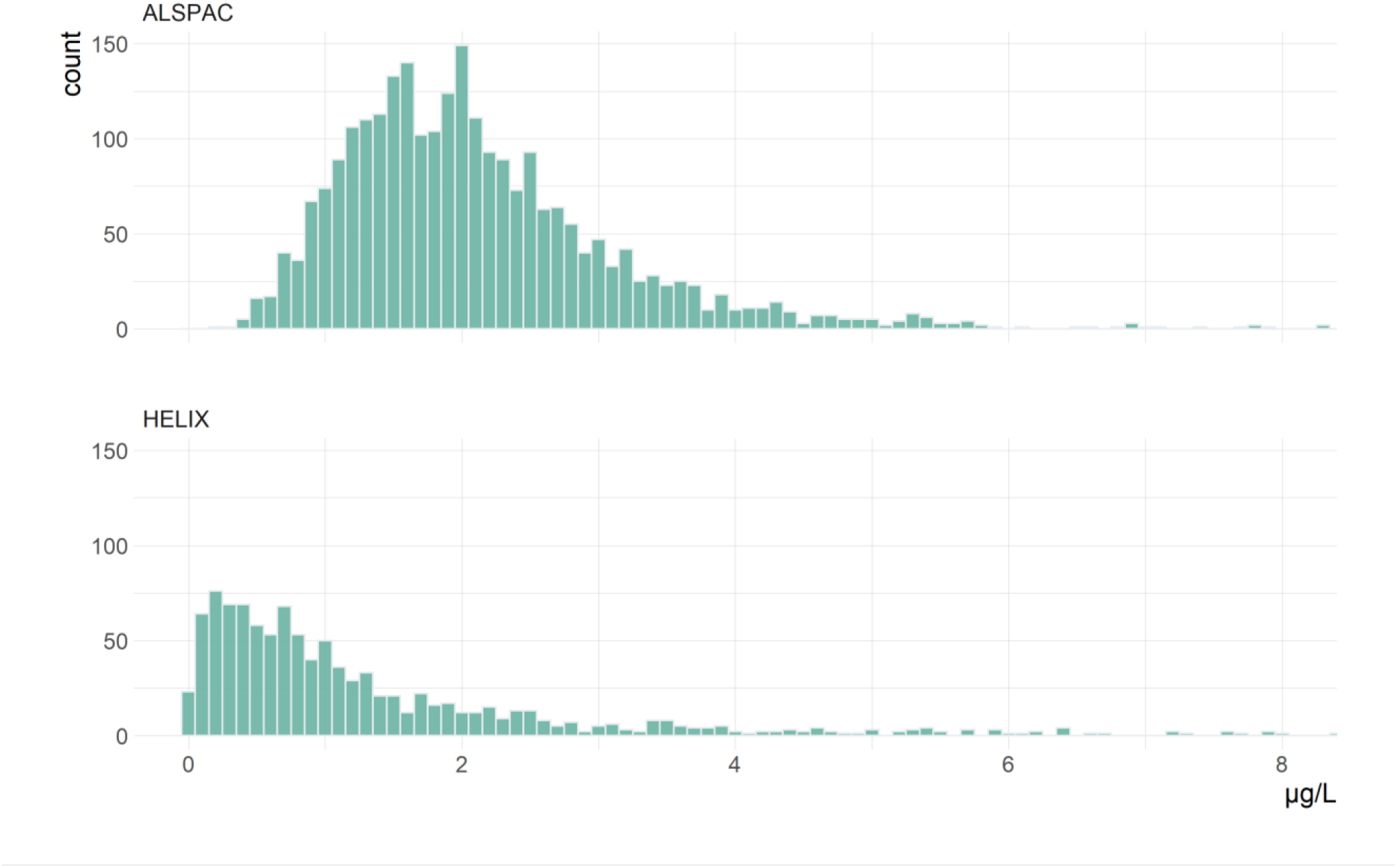
Blood mercury concentrations in (a) 2,893 pregnant women in ALSPAC and (b) 1,042 children aged 6-11 years old in HELIX. Samples with Hg > 8 μg/L are excluded for readability (n=7, Figure S1 for same figure with no exclusions).

### SNP-heritability

Genome-wide SNP heritability (h^2^g) of Hg was calculated from GWAS summary statistics. There were summary statistics available for 6,620,135 SNPs from 2,893 pregnant women, and 6,138,843 SNPs for 1,042 children. These were regressed on LD Scores from the 1000 Genome Project Europeans reference panel. After merging with the reference panel, the LD Score regression included 1,137,154 and 965,135 SNPs for pregnant women and children, respectively.

The estimated h^2^g for pregnant women was 24.0% (95% confidence interval = 16.9% to 46.4%, p = 0.01). For children, the estimated was 4.8% but this was not statistically significant (p = 0.85) and confidence intervals overlapped with zero (95% CI: -45.7% to 55.4%).

### Genome-wide association studies

Manhattan plots summarising GWAS results in each population, and QQ plots of expected and observed p-values are shown in Supplementary Figures S3-4. There did not appear to be notable p-value inflation or deflation in QQ plots, or their lambda statistics which were 1.01 for pregnant women and 1.00 for children.

For pregnant women no genetic locations were found which were associated at genome-wide significant levels with Hg (P < 5×10^−8^), but 16 independent loci were found with at least 1 variant suggestively significant (P < 1×10^−5^). Similarly for children no genome-wide significant locations were found but there were 21 independent genetic loci which were suggestively associated with Hg. Summary statistics of suggestively significant variants are presented in Table 2.

**Table 2.** Summary statistics for the variants suggestively associated (p < 1×10^−5^) with blood Hg concentrations, pruned to the most significant SNP per independent genetic loci.

| **Pregnant women, ALSPAC (n = 2,893, nSNP = 6,620,135)** |  |  |  |  |  |  |  |  |  |  |  |
| --- | --- | --- | --- | --- | --- | --- | --- | --- | --- | --- | --- |
| SNP | Chr | Position (GRCH37) | Gene | Effect allele | Other allele | MAF | Beta | SE | P-value | HELIX beta | HELIX P-value |
| rs4853739 | 2 | 191698516 | - | T | C | 0.24 | 0.10 | 0.02 | $2.32 \times 10^{-06}$ | - | - |
| rs11709754 | 3 | 149615531 | *RNF13* | T | A | 0.21 | -0.10 | 0.02 | $9.39 \times 10^{-06}$ | - | - |
| rs361166 | 4 | 152792791 | - | A | G | 0.41 | 0.09 | 0.02 | $5.78 \times 10^{-06}$ | 0.05 | 0.53 |
| rs6859392 | 5 | 63241481 | - | G | C | 0.30 | -0.09 | 0.02 | $3.31 \times 10^{-06}$ | - | - |
| rs1372504 | 5 | 103749428 | *RP11-6N13.1* | A | G | 0.38 | -0.09 | 0.02 | $1.70 \times 10^{-06}$ | 0.06 | 0.53 |
| rs2246509 | 6 | 156058999 | - | G | A | 0.46 | 0.08 | 0.02 | $8.69 \times 10^{-06}$ | 0.00 | 0.98 |
| rs1845418 | 7 | 24095415 | - | T | C | 0.14 | 0.14 | 0.03 | $8.68 \times 10^{-08}$ | -0.09 | 0.48 |
| rs146099921 | 8 | 22254947 | *SLC39A14* | G | T | 0.14 | -0.12 | 0.03 | $8.21 \times 10^{-06}$ | 0.19 | 0.14 |
| rs7900717 | 10 | 122034207 | - | C | T | 0.25 | -0.09 | 0.02 | $6.02 \times 10^{-06}$ | -0.21 | 0.31 |
| rs7301395 | 12 | 116402976 | *MED13L* | A | G | 0.02 | 0.33 | 0.07 | $5.06 \times 10^{-06}$ | - | - |
| rs12874443 | 13 | 38304884 | *TRPC4* | T | G | 0.45 | -0.08 | 0.02 | $7.73 \times 10^{-06}$ | 0.00 | 0.99 |
| rs113202356 | 16 | 27891845 | *GSG1L* | A | G | 0.10 | -0.14 | 0.03 | $8.45 \times 10^{-06}$ | - | - |
| rs74450576 | 16 | 73918519 | - | A | G | 0.04 | 0.21 | 0.05 | $7.40 \times 10^{-06}$ | -0.05 | 0.80 |
| rs11643897 | 16 | 78221370 | *WWOX* | C | T | 0.32 | 0.08 | 0.02 | $9.70 \times 10^{-06}$ | 0.02 | 0.87 |
| rs35522803 | 19 | 3592734 | *GIPC3* | T | C | 0.08 | -0.15 | 0.03 | $3.83 \times 10^{-06}$ | 0.04 | 0.82 |
| rs60192794 | 20 | 52317186 | - | C | T | 0.07 | -0.16 | 0.04 | $5.66 \times 10^{-06}$ | - | - |
| **Children, HELIX (n = 1,042, nSNP = 6,138,843)** |  |  |  |  |  |  |  |  |  |  |  |
| SNP | Chr | Position (GRCH37) | Gene | Effect allele | Other allele | MAF | Beta | SE | P-value | ALSPA C beta | ALSPAC P-value |
| rs7526817 | 1 | 28195486 | - | A | T | 0.09 | -0.67 | 0.15 | $5.02 \times 10^{-06}$ | 0.06 | 0.06 |
| rs79810835 | 1 | 146999434 | - | A | G | 0.02 | 1.37 | 0.31 | $6.83 \times 10^{-06}$ | - | - |
| rs59436870 | 2 | 101830050 | TBC1D8 | T | C | 0.18 | -0.52 | 0.11 | 5.16x10<sup>-06</sup> | 0.00 | 0.74 |
| rs9852537 | 3 | 98658085 | CTD-2021J15.1 | T | C | 0.03 | 1.29 | 0.29 | 7.62x10<sup>-06</sup> | 0.02 | 0.66 |
| rs186276942 | 3 | 115279279 | - | G | A | 0.01 | 2.87 | 0.63 | 5.29x10<sup>-06</sup> | - | - |
| rs62287513 | 3 | 184104050 | CHRD | A | G | 0.31 | 0.44 | 0.09 | 7.69x10<sup>-07</sup> | - | - |
| rs28618224 | 4 | 21041710 | KCNIP4 | C | A | 0.03 | 1.47 | 0.32 | 5.03x10<sup>-06</sup> | 0.09 | 0.11 |
| rs115812569 | 4 | 42726300 | - | G | A | 0.01 | 1.82 | 0.40 | 6.42x10<sup>-06</sup> | -0.04 | 0.56 |
| rs2904271 | 4 | 90284516 | - | C | A | 0.37 | -0.41 | 0.09 | 1.84x10<sup>-06</sup> | 0.00 | 0.79 |
| rs113384484 | 5 | 151015719 | - | A | G | 0.04 | 1.03 | 0.22 | 1.50x10<sup>-06</sup> | 0.11 | 0.02 |
| rs79340261 | 7 | 35559836 | - | A | C | 0.02 | 1.93 | 0.40 | 1.08x10<sup>-06</sup> | - | - |
| rs75847252 | 9 | 71483481 | PIP5K1B | C | T | 0.02 | 1.22 | 0.26 | 2.96x10<sup>-06</sup> | - | - |
| rs73510541 | 11 | 69313573 | - | A | G | 0.13 | 0.54 | 0.12 | 3.72x10<sup>-06</sup> | 0.02 | 0.50 |
| rs9510838 | 13 | 24317031 | - | T | G | 0.17 | 0.58 | 0.12 | 3.43x10<sup>-06</sup> | -0.00 | 0.88 |
| rs9563673 | 13 | 34239526 | RP11-141M1.3 | C | T | 0.07 | -0.90 | 0.19 | 5.90x10<sup>-06</sup> | 0.05 | 0.21 |
| rs17106291 | 14 | 37559401 | SLC25A21 | G | A | 0.02 | 2.32 | 0.52 | 7.53x10<sup>-06</sup> | - | - |
| rs7154700 | 14 | 63201771 | KCNH5 | T | C | 0.01 | 1.61 | 0.34 | 2.15x10<sup>-06</sup> | - | - |
| rs116971963 | 17 | 14961821 | - | A | G | 0.01 | 1.57 | 0.34 | 4.91x10<sup>-06</sup> | - | - |
| rs145982353 | 19 | 54257385 | - | G | C | 0.03 | 1.27 | 0.27 | 4.07x10<sup>-06</sup> | - | - |
| rs6075980 | 20 | 288233 | - | G | A | 0.38 | 0.42 | 0.09 | 2.31x10<sup>-06</sup> | -0.02 | 0.24 |
| rs148653405 | 20 | 5594216 | - | A | G | 0.01 | 1.77 | 0.40 | 8.35x10<sup>-06</sup> | -0.06 | 0.34 |
| “-“ indicates the variant was not available in the other study. |  |  |  |  |  |  |  |  |  |  |  |
| Variants annotated to genes only if the SNP is in the gene body. |  |  |  |  |  |  |  |  |  |  |  |
| MAF: Minor allele frequency |  |  |  |  |  |  |  |  |  |  |  |
| SE: Standard error |  |  |  |  |  |  |  |  |  |  |  |

None of the 37 suggestively associated SNPs were also associated with Hg in the other population even at a more relaxed p-value threshold of p >1×10^−2^. Results for 15 of these SNPs were not available in the other study, including 9 which were low frequency (MAF < 5%) and likely excluded during earlier stages of the analysis.

### In silico functional analysis

37 SNPs which were suggestively significant in the two GWAS were mapped to the nearest genes using NCBI Sequence Viewer and FUMA SNP2Gene (Supplementary Table S4). Additionally, we identified variants associated with gene expression (P > 5×10^−8^) using Phenoscanner, and included these genes in the following analyses.

SNPs were most commonly associated with gene expression, histone modification and methylation at genes or CpG sites close to the SNP locations. No SNP or variants in strong LD (r^2^>0.8) were found to have known links to Hg metabolism. Results for each SNP are summarised in Supplementary Table S5.

In ALSPAC the T allele of intronic variant rs146099921 was associated with -0.12 log Hg (p = 8.21×10^−6^). This variant is in the gene Solute Carrier Family 39 Member 14 *(SLC39A14)*. This variant was associated with reduced DNA methylation at cg14348540 in *SLC39A14* (90, 91), and exon expression in *SLC39A14* (92). The gene is a metal transporter linked to cellular uptake of cadmium, iron, manganese, and zinc, and also referred to as *ZIP14* (93). Mutations are associated with the impairment of manganese transport and homeostasis, leading to toxic accumulation (94-96). There is evidence that the gene also functions as a transporter of zinc (97), and mediates iron and cadmium uptake (98, 99), with a detailed review of metal transport functions available elsewhere (93). According to data available in GTEx Portal (88), the gene is expressed throughout the body but most highly in the liver followed by the adipose tissue, the arteries, and pancreas.

There were further links between suggestive variants and genes with functions potentially affecting Hg levels, such as for rs17106291 (Solute Carrier Family 25 Member 21, *SLC25A21*) which transports dicarboxylates across the inner membranes of mitochondria (100). Two variants (rs28618224, rs7154700) were in potassium voltage-gated channels genes, and two variants were near to genes affecting glutamate (rs361166) and phospholipid (rs115812569) transport.

### Associations in previous candidate variants

GWAS summary statistics were extracted for 14 variants which were identified in previous genetic association studies or GWAS of metals that may interact with mercury (Supplementary Table S2). These variants are reported in Table 3 with summary statistics from ALSPAC and HELIX GWAS. The minor C allele of rs10636 was associated with increased blood Hg in pregnant women (p=0.01), and the minor C allele rs9936741 was associated with lower Hg in children (p = 0.01). Neither variant showed strong evidence of association in the alternate GWAS, and no association was found in variants previously reported as significant in GWAS of blood lead, selenium, or zinc (Table 3).

**Table 3.**
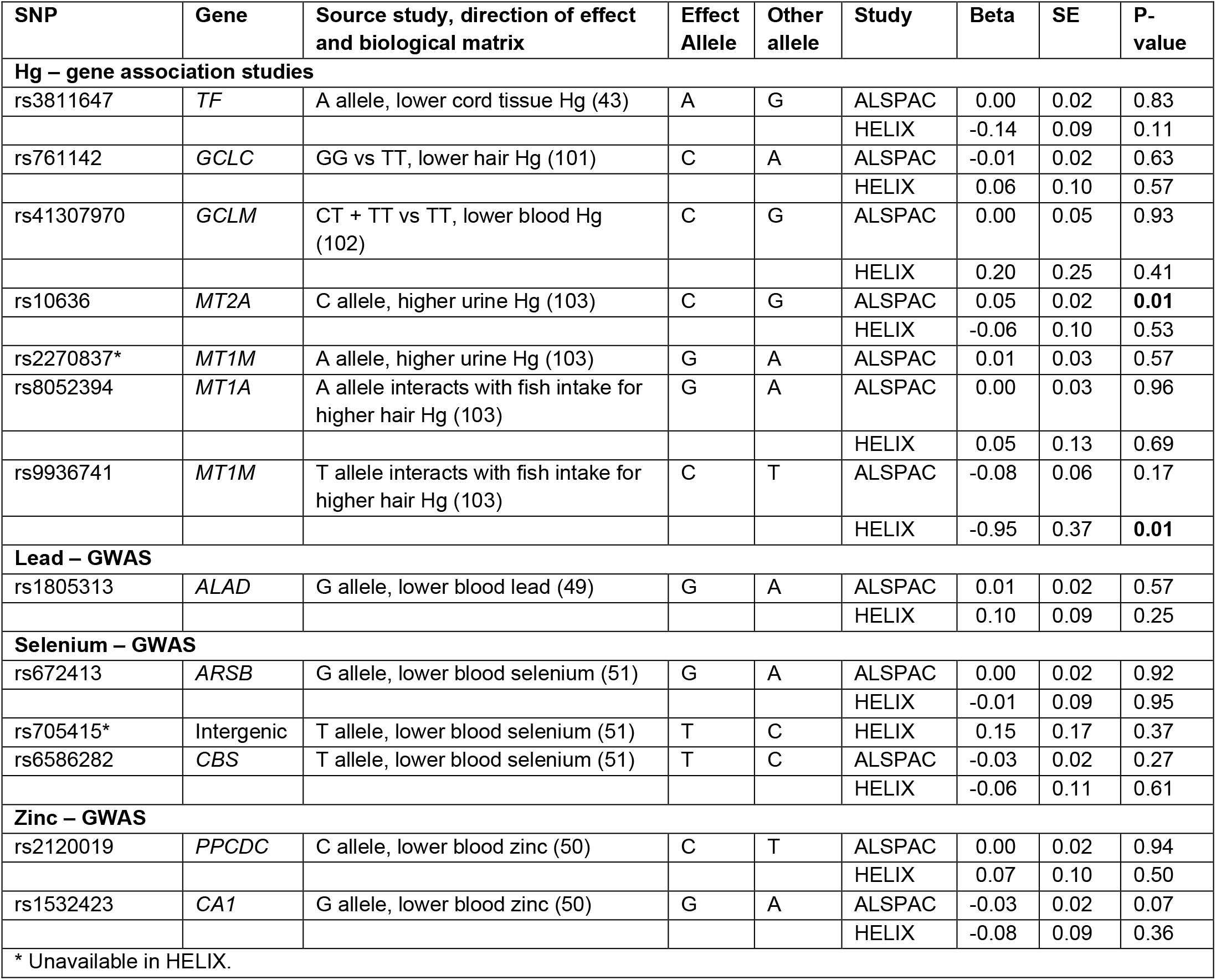
SNP-Hg summary statistics for candidate variants identified in previous studies of Hg or other metal GWAS.

| SNP | Gene | Source study, direction of effect and biological matrix | Effect Allele | Other allele | Study | Beta | SE | P-value |
| --- | --- | --- | --- | --- | --- | --- | --- | --- |
| **Hg – gene association studies** |  |  |  |  |  |  |  |  |
| rs3811647 | *TF* | A allele, lower cord tissue Hg (43) | A | G | ALSPAC | 0.00 | 0.02 | 0.83 |
|  |  |  |  |  | HELIX | -0.14 | 0.09 | 0.11 |
| rs761142 | *GCLC* | GG vs TT, lower hair Hg (101) | C | A | ALSPAC | -0.01 | 0.02 | 0.63 |
|  |  |  |  |  | HELIX | 0.06 | 0.10 | 0.57 |
| rs41307970 | *GCLM* | CT + TT vs TT, lower blood Hg (102) | C | G | ALSPAC | 0.00 | 0.05 | 0.93 |
|  |  |  |  |  | HELIX | 0.20 | 0.25 | 0.41 |
| rs10636 | *MT2A* | C allele, higher urine Hg (103) | C | G | ALSPAC | 0.05 | 0.02 | **0.01** |
|  |  |  |  |  | HELIX | -0.06 | 0.10 | 0.53 |
| rs2270837* | *MT1M* | A allele, higher urine Hg (103) | G | A | ALSPAC | 0.01 | 0.03 | 0.57 |
| rs8052394 | *MT1A* | A allele interacts with fish intake for higher hair Hg (103) | G | A | ALSPAC | 0.00 | 0.03 | 0.96 |
|  |  |  |  |  | HELIX | 0.05 | 0.13 | 0.69 |
| rs9936741 | *MT1M* | T allele interacts with fish intake for higher hair Hg (103) | C | T | ALSPAC | -0.08 | 0.06 | 0.17 |
|  |  |  |  |  | HELIX | -0.95 | 0.37 | **0.01** |
| **Lead – GWAS** |  |  |  |  |  |  |  |  |
| rs1805313 | *ALAD* | G allele, lower blood lead (49) | G | A | ALSPAC | 0.01 | 0.02 | 0.57 |
|  |  |  |  |  | HELIX | 0.10 | 0.09 | 0.25 |
| **Selenium – GWAS** |  |  |  |  |  |  |  |  |
| rs672413 | *ARSB* | G allele, lower blood selenium (51) | G | A | ALSPAC | 0.00 | 0.02 | 0.92 |
|  |  |  |  |  | HELIX | -0.01 | 0.09 | 0.95 |
| rs705415* | Intergenic | T allele, lower blood selenium (51) | T | C | HELIX | 0.15 | 0.17 | 0.37 |
| rs6586282 | *CBS* | T allele, lower blood selenium (51) | T | C | ALSPAC | -0.03 | 0.02 | 0.27 |
|  |  |  |  |  | HELIX | -0.06 | 0.11 | 0.61 |
| **Zinc – GWAS** |  |  |  |  |  |  |  |  |
| rs2120019 | *PPCDC* | C allele, lower blood zinc (50) | C | T | ALSPAC | 0.00 | 0.02 | 0.94 |
|  |  |  |  |  | HELIX | 0.07 | 0.10 | 0.50 |
| rs1532423 | *CA1* | G allele, lower blood zinc (50) | G | A | ALSPAC | -0.03 | 0.02 | 0.07 |
|  |  |  |  |  | HELIX | -0.08 | 0.09 | 0.36 |
| * Unavailable in HELIX. |  |  |  |  |  |  |  |  |

## Discussion

Genome-wide association analyses of blood Hg in British pregnant women and in children from six European cohorts did not find robust associations with any genetic variants. Despite no single variant being strongly linked to Hg, genome-wide SNP heritability was estimated to explain a considerable proportion of Hg variance in pregnant women (24.0%, 95% CI: 16.9 to 46.4). Considering that Hg is highly reactive with a wide range of molecules and exposure is affected by numerous biological processes, we anticipated a substantial component of its metabolism would be heritable and our finding is consistent with animal and plant studies (104, 105) which estimated large genetic components of phenotypic variation.

Although no variants passed genome-wide significance thresholds, there were 37 independent loci detected in the two studies with SNPs significant at a lower suggestive threshold. Yet these results were in low concordance between pregnant women and children, with several possible reasons. First, differences may be due to qualitative differences in Hg metabolism between the two populations if metabolism changes with age. This does not appear to have been studied in human populations, but animal studies have reported changes in early-life absorption and excretion (106, 107).Secondly, the analyses may have been underpowered, which would restrict the ability for the GWAS to estimate true associations. The sample of children was considerably smaller and had lower median Hg levels than pregnant women, and in our heritability analysis there was very wide uncertainty (h^2^g = 4.8%,95% CI: -45.7% to 55.4%) which is indicative of low power. Finally, if there are non-linear associations between variants and Hg this may have contributed to the heterogeneity we found, because as seen in Figure 1, Hg distributions were not identical.

From the suggestive variants, the most biologically plausible link to Hg was with rs146099921 in gene *SLC39A14*. The variant modifies expression of *SLC39A14* (92), and local methylation (90, 91) and exon expression (92). Most SLC39 genes are responsible for the cellular uptake of zinc (108), but studies suggest *SLC39A14*, also referred to as *ZIP14*, is associated with changes in cadmium, iron, and manganese levels (93, 94, 98). It is possible the gene impacts Hg levels indirectly through these metals, each of which may interact with Hg - for example increased cellular zinc induces metallothionein synthesis and may increase removal of Hg (109). Alternatively, the gene may directly affect the transport of Hg, but this does not appear to be reported in prior studies. In children the variant rs17106291 in another SLC gene (*SLC25A21*) was also related to Hg levels at suggestive significance, which is a transport of C5-C7 oxodicarboxylates to mitochondria. Other members of the *SLC* gene family have been linked to kidney uptake of I-Hg (110) and the intestinal transport of MeHg (15).

Several other suggestively significant SNPs were annotated to genes with possible connections to Hg metabolism. In children, variants were located in potassium voltage-gated channels genes *KCNH5* and *KCNIP4*, in ALSPAC a calcium ion channel gene *TRPC4*. The expression of MeHg and I-Hg toxicity may be linked to inhibited potassium or calcium channels (109), although it is unclear how variation in these genes would impact Hg blood levels. Finally, in pregnant women there were variants located near to genes affecting glutamate (*GLS, GATB*). Hg both inhibits glutamate uptake (111) and stimulates its release (112).

This study did not replicate many of the findings reported in previous gene association studies of Hg. From the literature, we identified 7 variants of interest to Hg metabolism, located in genes *GCLM, GCLC, TF, MT1A, MT1M*, and *MT2A*. In both pregnant women and children, most variants were not significantly associated with blood Hg, with two exceptions.

First, in pregnant women the variant rs10636 (*MT2A*) was nominally associated with Hg in the same direction as in a previous study (103). Secondly, in children we found that rs9936741 (*MT1M*) was found to be associated with child Hg in the opposite direction to that reported previously (41), which could be due to the use of hair Hg and an interaction term with diet in the original study. There is a biologically plausible role for these variants, as metallothionein genes generate proteins which bind to Hg and aid clearance (113). However, neither result was replicated in both cohorts and multiple testing may have increased the likelihood of finding variants with low p-values.

The overall lack of replication between this study and previous gene association studies may be because of the smaller sample sizes used in the earlier candidate gene studies (Supplementary Table S2 for details). Finally, an epigenome-wide analysis of umbilical cord Hg identified associations in the genes *GGH, MED31*, and *GRK1* and DNA methylation (114). The direction of causality is unclear, but in this study no strong signals were found in variants near the reported CpG sites.

Limitations of this study were as follows. First, the lack of genome-wide significant SNPs and high heterogeneity between studies suggests the analyses may have been underpowered. By comparison, larger sized GWAS identified one loci associated with blood lead levels (n=5,433) (49) and two with selenium levels (n=9,639) (51). The required sample size will also vary by the variation of the trait, as demonstrated by an arsenic GWAS in Bangladesh which was able to identify five independent loci from 1,313 arsenic-exposed individuals (115). The low power may be part of the reason for the discordance between ALSPAC and HELIX results, and in particular for SNPs with low MAF.

A second caveat of this study is that blood Hg is primarily in the form of MeHg, with a smaller proportion possibly as I-Hg (116, 117). Blood Hg reflects relatively short-term exposure and therefore is subject to day-to-day variation, which increases residual error in linear models and reduces study power. Other biological matrices such as hair or nail Hg may be more representative of long-term Hg exposure and therefore avoid this issue, and this is something future studies should consider.

Third, the HELIX study comprised 6 subcohorts located in different countries. These populations will have different patterns of Hg exposure from dietary and environmental variation, and this may have introduced heterogeneity into the pooled analysis. Finally, this GWAS was conducted on a population that is mostly of European ancestry, and where exposure to Hg is likely to be primarily through fish consumption. Populations with different ancestries or where there are different sources of exposure such as local environmental exposure may have different genotype-Hg associations.

## Conclusion

This study found that SNP-heritability was associated with a considerable amount of Hg variance in pregnant women. No genome-wide significant variants were found, but rs146099921 was located in a transporter of multiple metals (*SLC39A14*) was suggestively associated with Hg levels in pregnant women. There was low correlation between results from pregnant women and children, which could reflect changes in Hg metabolism during development. The study is continuing to seek collaboration with other institutes that may have comparable populations, to increase the quantity and accuracy of variants identified.

## Data Availability

GWAS summary statistics for LD clumped SNPs which were below a suggestive p-value threshold are reported in Table 2. Full summary statistics may be released at a later date following final publication of the study.

## Acknowledgements

We are extremely grateful to all the families who took part in the ALSPAC and HELIX studies, the midwives for their help in recruiting them, and the whole ALSPAC and HELIX teams, which includes interviewers, computer and laboratory technicians, clerical workers, research scientists, volunteers, managers, receptionists and nurses.

## Ethics approval

Ethics approval for the study was obtained from the ALSPAC Ethics and Law Committee and the Local Research Ethics Committees. Consent for biological samples has been collected in accordance with the Human Tissue Act (2004). Informed consent for the use of data collected via questionnaires and clinics was obtained from participants following the recommendations of the ALSPAC Ethics and Law Committee at the time.

In HELIX, local ethical committees approved the studies that were conducted according to the guidelines laid down in the Declaration of Helsinki. The ethical committees for each cohort were the following: BIB: Bradford Teaching Hospitals NHS Foundation Trust, EDEN: Agence nationale de sécurité du médicament et des produits de santé, INMA: Comité Ético de Inverticación Clínica Parc de Salut MAR, KANC: LIETUVOS BIOETIKOS KOMITETAS, MoBa: Regional komité for medisinsk og helsefaglig forskningsetikk, Rhea: Ethical committee of the general university hospital of Heraklion, Crete. Informed consent was obtained from a parent and/or legal guardian of all participants in the study. Participants did not receive any compensation.

