## Supplementary Materials for "Genome-wide association study of blood mercury in European pregnant women and children"

**Supplementary file**

**Supplementary Table S1.** Summary of study population, measurement methods, and GWAS characteristics.

|  | **ALSPAC** | **HELIX** |
| --- | --- | --- |
| Date of recruitment | April 1991 – December 1992 | 1999 - 2010 |
| Population | Pregnant women | Children aged 7 to 9 |
| Location | Avon Health Authority area, UK | UK, France, Spain, Lithuania, Norway, and Greece |
| N recruited | 14,833 | Approximately 32,000 |
| Mercury sample source | Whole blood | Whole blood |
| Mercury sample timing | Early pregnancy | 6 – 11 years old |
| Mercury sample analysis method | ICP-DRC-MS | ICP-SFMS |
| N with mercury sample | 4,131 | 1,301 |
| **Genotyping** |  |  |
| N with blood DNA sample | 10,015 | 1,397 |
| Genotyping method | Illumina Human660W-Quad Array | Infinium Global Screening Array (GSA) (Illumina) |
| N SNP directly genotyped | 557,124 | 692,367 |
| QC software | Plink v1.07 | Plink v1 |
| Individual missingness | >5% | >3% |
| SNP missingness | >5% | >5% |
| Hardy-Weinberg equilibrium | P < 1.0x10^-07^ | P < 1.0x10^-06^ |
| Minor allele frequency | <1% | <1% |
| Cryptic relatedness | IBD > 0.125 | IBD > 0.125 |
| Other exclusions | indeterminate X chromosome heterozygocity, extreme autosomal heterozygocity, population outliers | Gender mismatch, heterozygosity, relatedness, non-canonical PAR |
| N samples post QC | 8,196 | 1,304 |
| N SNP post QC | 526,688 | 509,344 |
| **Imputation** |  |  |
| Reference panel | Haplotype Reference Consortium (HRC r1.1) | Haplotype Reference Consortium (HRC r1.1) |
| Phasing | ShapeIt v2 | Eagle v2.4 |
| Imputation | Impute V3 | minimac4 |
| N SNP after imputation | 39,117,141 | 40,405,505 |
| Minor allele frequency | <1% | <1% |
| Imputation quality (INFO) | <0.9 | <0.9 |
| Hardy-Weinberg equilibrium | P < 1.0x10^-07^ | P < 1.0x10^-06^ |
| N SNP post QC | 6,649,782 | 6,143,757 |
| **GWAS** |  |  |
| N samples mercury + genotype | 2,893 | 1,042 |
| GWAS software | SNPTEST 2.5.2 | Plink v1 |
| Covariates | Age + 10 PCA | Age + 10 PCA |
| Minor allele frequency | <1% | <1% |
| Other exclusions | Unidentifiable rsid | Unidentifiable rsid |

**Supplementary Table S2**

1. Variants identified in gene-association studies of mercury.

| **SNP** | **Gene** | **Biological sample** | **Population** | **Effect allele** | **Other allele** | **Beta (p-value)** | **PMID** |
| --- | --- | --- | --- | --- | --- | --- | --- |
| rs41307970 | *GCLM* | Whole blood & plasma | 88 adults in Amazon riverside communities. | CT + TT | CC | −0.21 (0.05) | 24696865 |
| rs2270837 | *MT1M* | Urine | 515 dental professionals | A | G | 1.85 (0.008) | 22233731 |
| rs10636 | *MT2A* | Urine | 515 dental professionals | C | G | 0.06 (-) | 22233731 |
| rs8052394 | *MT1A* | Hair | 515 dental professionals | A * intake | G | -300 (0.02) | 22233731 |
| rs9936741 | *MT1M* | Hair | 515 dental professionals | T * intake | C | 19.3 (0.02) | 22233731 |
| rs761142 | *GCLC* | Hair | 1,449 mothers in Seychellois population | GG | TT | -0.46 (0.02) | 29573653 |
| rs3811647 | *TF* | Cord tissue | 1,311 cord samples | A | G | - (0.03) | 23903878 |

2. Variants identified in genome-wide association studies of metals that interact with mercury within the body (lead, selenium, zinc).

| **SNP** | **Gene** | **Metal** | **Population** | **Effect allele** | **Other allele** | **Beta (p-value)** | **PMID** |
| --- | --- | --- | --- | --- | --- | --- | --- |
| rs1805313 | *ALAD* | Lead | 5,433 Australian and UK adults. | A | G | + (3.91 × 10^−14^) | 25820613 |
| rs672413 | *ARSB* | Selenium | 9,639 subjects from meta-analysis of US, Australian, and UK adults | A | G | + (5.21 x 10^−14^) | 25343990 |
| rs705415 | *-* | Selenium | 8,054 subjects from meta-analysis of US, Australian, and UK adults | T | C | - (4.64 x 10^−10^) | 25343990 |
| rs6586282 | *CBS* | Selenium | 9,639 subjects from meta-analysis of US, Australian, and UK adults | T | C | - (3.96 x 10^−09^) | 25343990 |
| rs1532423 | *CA1* | Zinc | 2,603 adults Australia | A | G | 0.18 (6.40 × 10^−12^) | 23720494 |
| rs2120019 | *PPCDC* | Zinc | 2,603 adults Australia | C | T | -0.29 (1.40 × 10^−12^) | 23720494 |

**Supplementary Table S3**

Summary of GWAS studies, number of partipants, and genetic variants.

| **Study** | **ALSPAC** | | **HELIX** | |
| --- | --- | --- | --- | --- |
| Direct genotyping | Illumina human660W quad | | Infinium Global Screening Array (GSA) (Illumina) | |
|  | **SNP** | **N** | **SNP** | **N** |
| Direct genotyped | 557,124 | 10,015 | 692,367 | 1,397 |
| Post QC | 526,688 | 8,196 | 509,344 | 1,304 |
| Imputation (HRC r1.1) | 39,117,141 | 8,196 | 40,405,505 | 1,304 |
| Post imputation QC | 6,649,782 | 8,196 | 6,143,757 | 1,304 |
| Mercury samples | - | 4,014 | - | 1,301 |
| GWAS | 6,649,782 | 2,893 | 6,143,757 | 1,042 |
| Post GWAS QC | 6,620,135 | 2,893 | 6,138,843 | 1,042 |

**Supplementary Figure S1**

Blood mercury concentrations in (a) 2,893 pregnant women in ALSPAC and (b) 1,042 children aged 6-11 years old in HELIX.

Includes 7 samples with Hg > 8 μg/L which were removed from Figure 1 for readability.

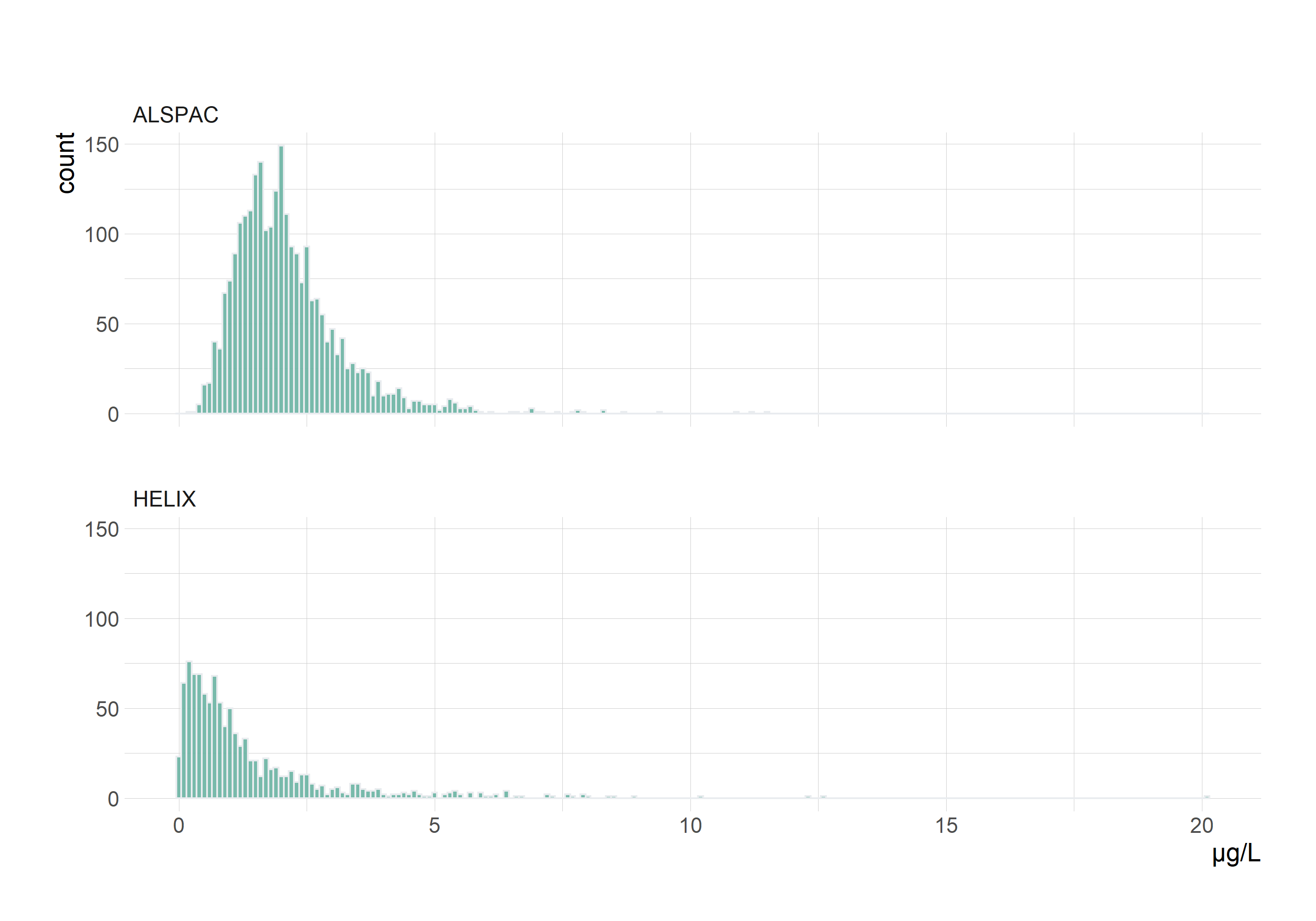

**Supplementary Figure S2**

Manhattan plot of GWAS results

(a) ALSPAC
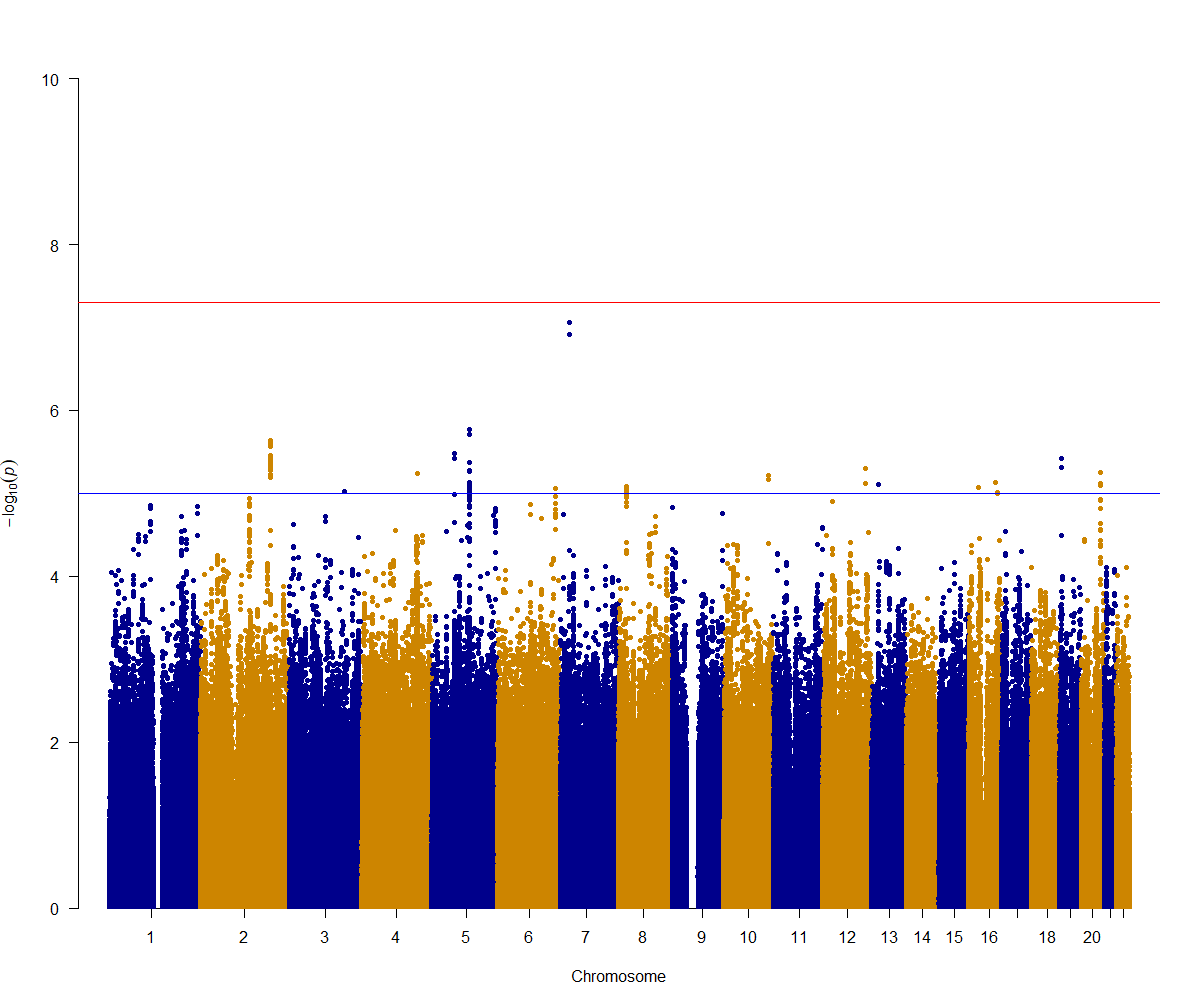

(b) HELIX
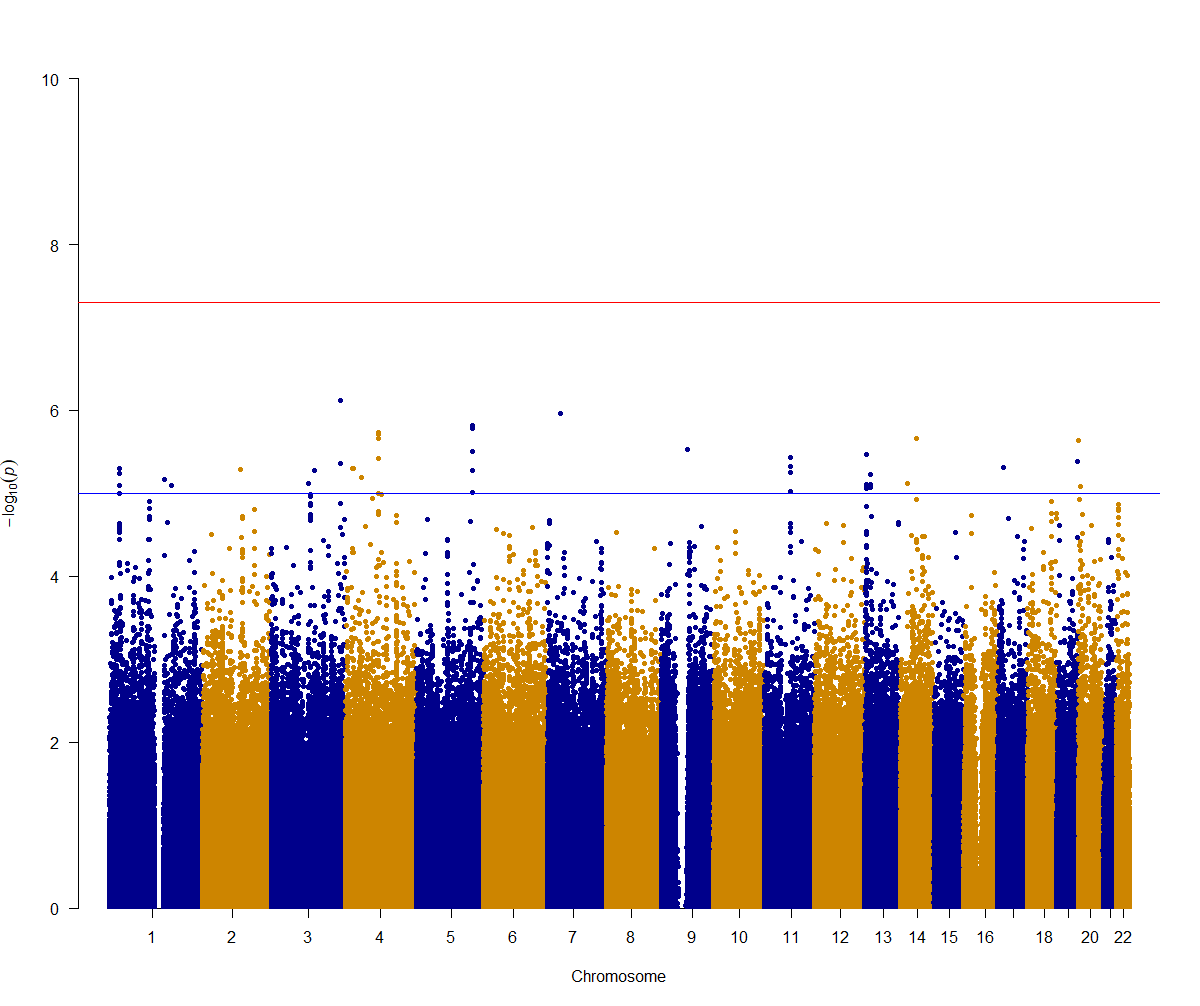

**Supplementary Figure S3**

QQ plot of expected and observed p-values.

(a) ALSPAC
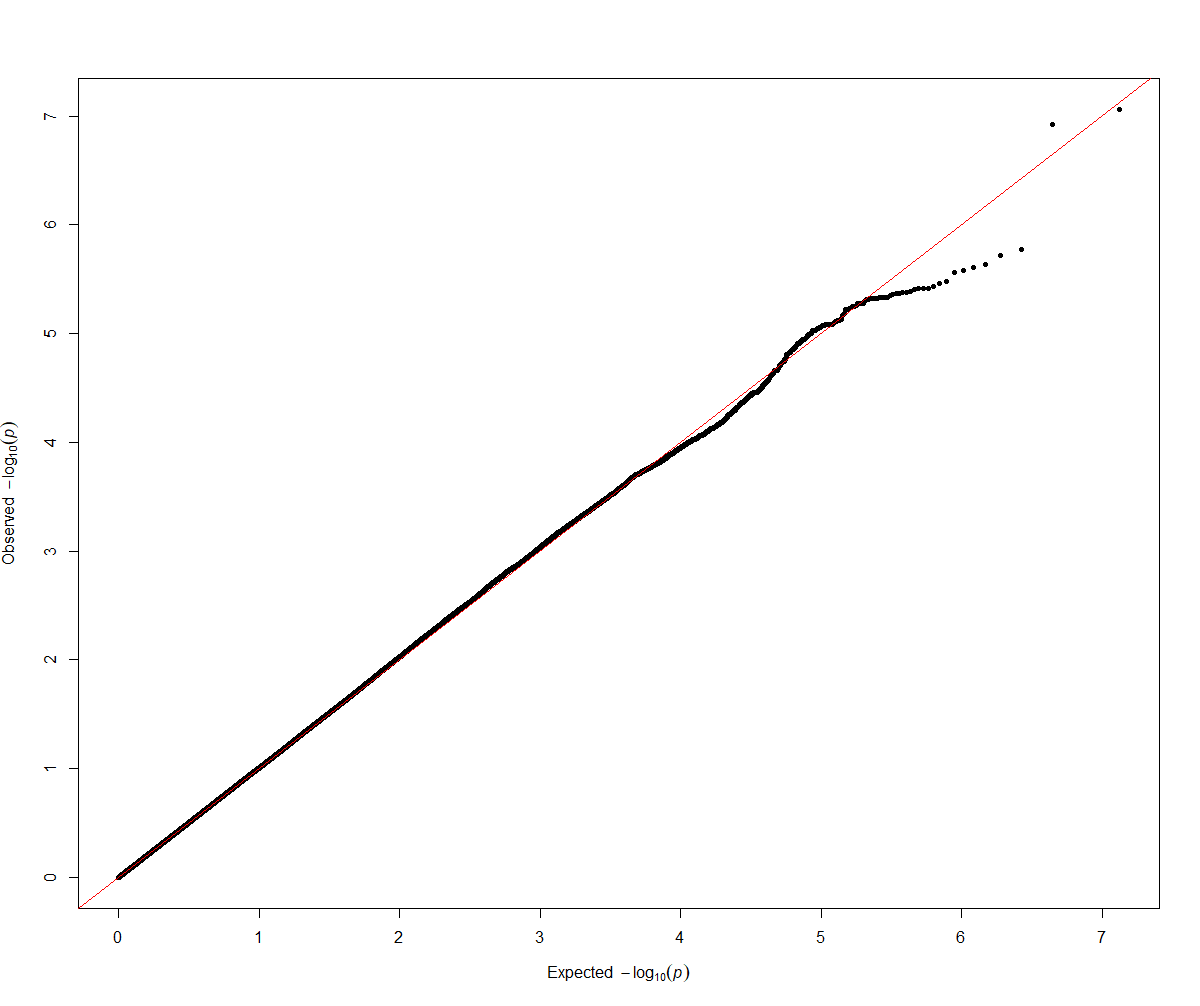

(b) HELIX
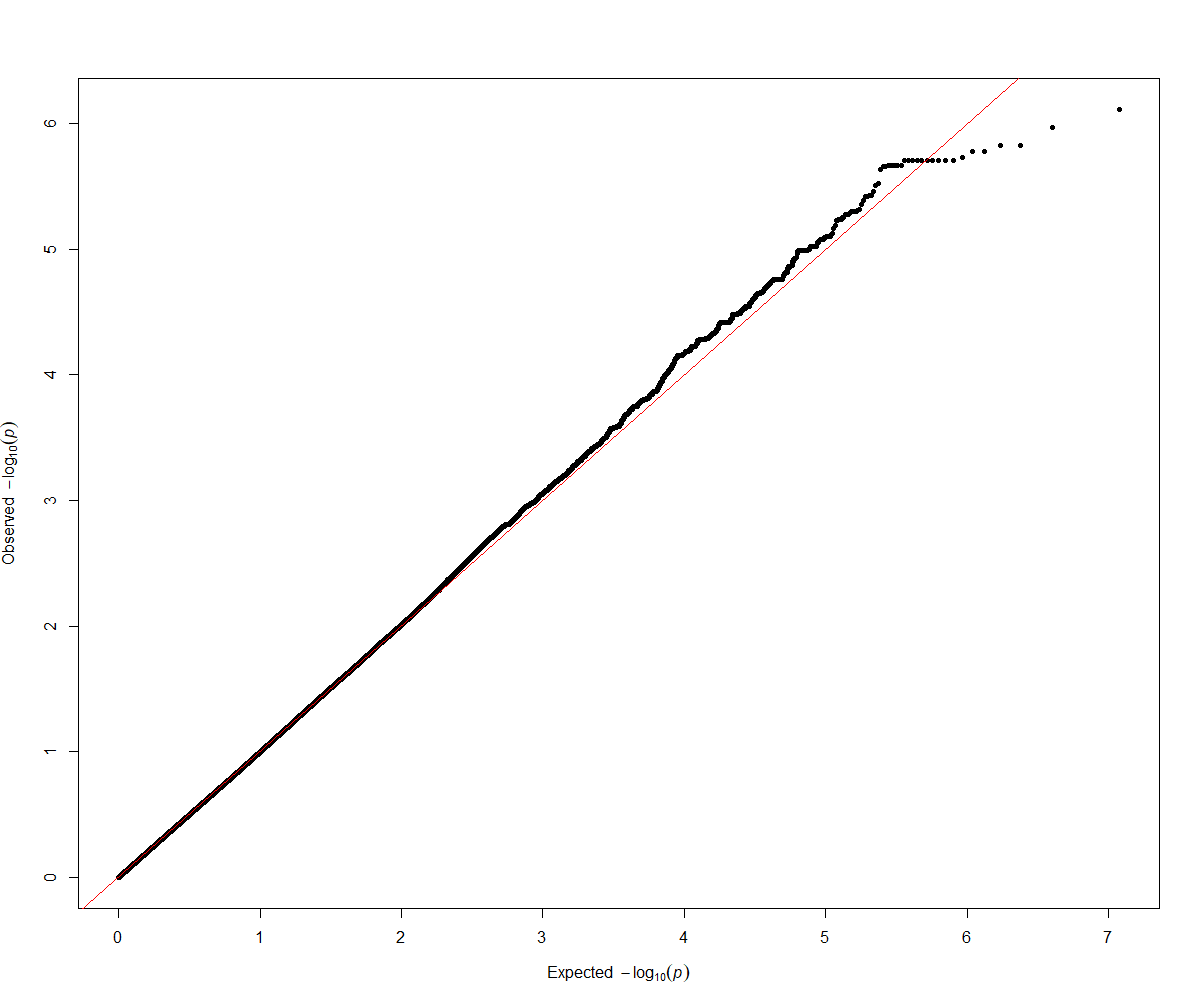

**Supplementary Table S4**

Gene mapping of LD clumped suggestive (p <1x10^-5^) variants. Automated mapping taken from FUMA SNP2Gene, and manually checked using dbSNP.

(a) ALSPAC

| **SNP** | **Chr** | **Position** (GRCH37) | **Gene** | **Distance to lead SNP** | **Type** |
| --- | --- | --- | --- | --- | --- |
| rs4853739 | 2 | 191698516 | *AC005540.3* | 47006 | intergenic |
| rs11709754 | 3 | 149615531 | *RNF13* | 0 | intronic |
| rs361166 | 4 | 152792791 | *GATB* | 110632 | Intergenic |
| rs6859392 | 5 | 63241481 | *RP11-158J3.2* | 12238 | Intergenic^1^ |
| rs1372504 | 5 | 103749428 | *RP11-6N13.1* | 0 | ncRNA_intronic |
| rs2246509 | 6 | 156058999 | *RNU7-152P* | 82838 | intergenic |
| rs1845418 | 7 | 24095415 | *RNA5SP228* | 76908 | intergenic |
| rs146099921 | 8 | 22248665 | *SLC39A14* | 0 | intronic |
| rs7900717 | 10 | 122034207 | *RPL21P16* | 79950 | intergenic |
| rs7301395 | 12 | 116396301 | *MED13L* | 0 | downstream |
| rs12874443 | 13 | 38303606 | *TRPC4* | 0 | intronic |
| rs113202356 | 16 | 27891845 | *GSG1L* | 0 | intronic |
| rs74450576 | 16 | 73918519 | *RPSAP56* | 56018 | intergenic |
| rs11643897 | 16 | 78221370 | *WWOX* | 0 | intronic |
| rs35522803 | 19 | 3592133 | *GIPC3* | 0 | UTR3 |
| rs60192794 | 20 | 52317186 | *RNU7-14P* | 31888 | intergenic |
| 1. In linkage disequilibrium with intronic variant within named gene. | | | | | |

(b) HELIX

| **SNP** | **Chr** | **Position** (GRCH37) | **Gene** | **Distance** | **Type** |
| --- | --- | --- | --- | --- | --- |
| rs7526817 | 1 | 28195486 | *THEMIS2* | 3568 | Intergenic^2^ |
| rs79810835 | 1 | 146999434 | *LINC00624* | 9734 | intergenic |
| rs59436870 | 2 | 101830050 | *TBC1D8* | 0 | intronic |
| rs9852537 | 3 | 98658085 | *CTD-2021J15.1* | 0 | ncRNA_intronic |
| rs186276942 | 3 | 115279279 | *GAP43* | 62891 | intergenic |
| rs62287513 | 3 | 184104050 | *CHRD* | 0 | intronic |
| rs28618224 | 4 | 21041710 | *KCNIP4* | 0 | intronic |
| rs115812569 | 4 | 42726300 | *ATP8A1* | 67178 | intergenic |
| rs2904271 | 4 | 90280428 | *GPRIN3* | 53437 | intergenic |
| rs113384484 | 5 | 151015719 | *CTB-113P19.4* | 16116 | intergenic |
| rs79340261 | 7 | 35559836 | *AC007652.1* | 9962 | intergenic |
| rs75847252 | 9 | 71483481 | *PIP5K1B* | 0 | intronic |
| rs73510541 | 11 | 69313573 | *AP000439.3* | 18864 | Intergenic^2^ |
| rs9510838 | 13 | 24300617 | *MIPEP* | 3710 | Intergenic^2^ |
| rs9563673 | 13 | 34236442 | *RP11-141M1.3* | 0 | ncRNA_intronic |
| rs17106291 | 14 | 37559401 | *SLC25A21* | 0 | intronic |
| rs7154700 | 14 | 63201771 | *KCNH5* | 0 | Intronic^1^ |
| rs116971963 | 17 | 14961821 | *AC005772.2* | 6227 | intergenic |
| rs145982353 | 19 | 54257385 | *MIR527* | 28 | downstream |
| rs6075980 | 20 | 288233 | *ZCCHC3* | 7267 | intergenic |
| rs148653405 | 20 | 5594216 | *GPCPD1* | 2543 | Upstream |
| 1. In linkage disequilibrium (r2>0.8) with UTR3 variant within named gene. | | | | | |
| 2. In linkage disequilibrium (r2>0.8) with intronic variant within named gene. | | | | | |

**Supplementary Table S5**

Summary of functional analysis for LD clumped suggestive (p< 1x10^-5^) variants.

GWAS Catalog: No results for any SNPs.

(a) ALSPAC

| **SNP** | **SNP-based search** | | | **Gene-based search (results for nearest gene unless otherwise stated)** | | | |
| --- | --- | --- | --- | --- | --- | --- | --- |
|  | **LDtrait** (r2>0.8) | **Phenoscanner** (p < 5 x 10^-8^) | **eQTL** | **Gene** | **GeneCards** | **GTEx** (blank: no notable variation) | **OMIM** (human or animal cells) |
| rs4853739 | - | *GLS* & *MFSD6* expression | *GLS* (-0.10, 7.22 x 10^-09^) | *AC005540.3^1^* | Glutamine hydrolysis catalyst. *MFSD6*: MHC class 1 binding. |  | - |
| rs11709754 | **-** | *PFN2* expression | *PFN2* (0.16, 4.10 x 10^-14^) | *RNF13* | Protein-protein interactions, specifics unknown | Brain, whole blood | - |
| rs361166 | Trans fatty acid levels | Histone modification, educational attainment | - | *GATB^1^* | Glutaminyl-tRNA synthetase, involved in glutamine transfer | Brain, heart, skin | - |
| rs6859392 | - | - | - | *RP11-158J3.2^2^* | - | Brain – hippocampus and front cortex | - |
| rs1372504 | Major depressive disorder, insomnia | Daytime napping, doctor visit for anxiety/depression. | - | *RP11-6N13.1* | - | Testis | - |
| rs2246509 | - | - | - | *RNU7-152P^1^* | - | Brain – amygdala | - |
| rs1845418 | - | - | - | *RNA5SP228^1^* | - | Kidneys | - |
| rs146099921 | - | - | *SLC39A14* (-0.16, 6.84 x 10^-09^) | *SLC39A14* | Metal transporter, cellular uptake of cadmium, iron, manganese, zinc | Arteries, liver, pancreas | Zinc importer ([Liuzzi et al, 2005](https://pubmed.ncbi.nlm.nih.gov/15863613/)) Elevated zinc transport ([Taylor et al, 2005](https://pubmed.ncbi.nlm.nih.gov/15642354/)) Iron uptake ([Liuzzi et al, 2006](https://pubmed.ncbi.nlm.nih.gov/16950869/), [Gao et al, 2008](https://pubmed.ncbi.nlm.nih.gov/18524764/)) Manganese transport ([Steimle et al, 2019](https://pubmed.ncbi.nlm.nih.gov/31699897/)) |
| rs7900717 | - | - | - | *RPL21P16^1^* | - | - | - |
| rs7301395 | - | - | - | *MED13L* | Mediator component between regulatory proteins and RNA transcription. | - | - |
| rs12874443 | - | - | - | *TRPC4* | Calcium ion channels | Arteries, uterus | - |
| rs113202356 | - | - | - | *GSG1L* | Synapse receptor activity | Arteries, brain | - |
| rs74450576 | - | - | - | *RPSAP56^1^* | - | - | - |
| rs11643897 | - | - | - | *WWOX* | Tumour suppression | - | - |
| rs35522803 | - | *GIPC3* expression | - | *GIPC3* | Hair bundle/cell maturation. | - | - |
| rs60192794 | - | - | - | *RNU7-14P^1^* | - | Testis | - |
| 1. Intergenic variant, gene listed is nearest. | | | | | | | |
| 2. Located outside of gene, and in linkage disequilibrium (r2>0.8) with variant within gene. | | | | | | | |

(b) HELIX

| **SNP** | **SNP-based search** | | | **Gene-based search (results for nearest gene unless otherwise stated)** | | | |
| --- | --- | --- | --- | --- | --- | --- | --- |
|  | **LDtrait** (r2>0.8) | **Phenoscanner** (p < 5 x 10^-8^) | **eQTL** | **Gene** | **GeneCards** | **GTEx** (blank: no notable variation) | **OMIM** (human or animal cells) |
| rs7526817 | **-** | *RPA2* expression | *THEMIS2* (0.9, 6.10 x 10^-10^) | *THEMIS2^2^* | T cell receptor signalling | Whole blood | - |
| rs79810835 | **-** | *ACP6* expression |  | *LINC00624^1^* | - | Testis | - |
| rs59436870 | **-** | *SNORD89* expression | *RNF19* (0.57, 3.15 x 10-28) | *TBC1D8* | GTPase activator activity, predicted to be involved in intracellular protein transport *SNORD89*: ovarian cancer progression.  *RNF19:* Ubiquitin ligase protein activity. | - | - |
| rs9852537 | - | Type 2 lactosamine alpha-2,3-sialyltransferase | - | *CTD-2021J15.1* | - | - | - |
| rs186276942 | - | - | - | *GAP43^1^* | Neuronal growth | Brain, nerve | - |
| rs62287513 | - | - | - | *CHRD* | Early vertebrate embryonic development | Ubiquitous – high in brain | - |
| rs28618224 | - | - | - | *KCNIP4* | Potassium voltage-gated channels | Brain | - |
| rs115812569 | - | - | *ATP8A1* (0.34, 8.29 x 10^-08^) | *ATP8A1^1^* | Lipid (phospholipids) transport across membranes | Brain, thyroid | - |
| rs2904271 | - | - | - | *GPRIN3^1^* | Neuron development. | Brain and lungs | - |
| rs113384484 | - | *SPARC* expression | - | *CTB-113P19.4^1^* | Bone collagen calcification. | Oesophagus, skin, brain. | - |
| rs79340261 | - | - | - | *AC007652.1^1^* | - | Testis | - |
| rs75847252 | - | *TJP2* expression. | - | *PIP5K1B* | Regulation of lipid messenger. |  | - |
| rs73510541 | - | - | - | *AP000439.3^1^* | - | Small intestines, colon, skin | - |
| rs9510838 | - | - | - | *MIPEP^1^* | Maturation of oxidative phosphorylation-related proteins | - | - |
| rs9563673 | - | - |  | *RP11-141M1.3* | - | - | - |
| rs17106291 | - | Treatment with antihypertensive | - | *SLC25A21* | Transports dicarboxylates across the inner membranes of mitochondria and participates in lysine, tryptophan, and hydroxylysine catabolism | Testis | - |
| rs7154700 | - | Cause of death: alcohol hepatitis. | - | *KCNH5* | Potassium voltage-gated channels | Brain | - |
| rs116971963 | - | - | - | *AC005772.2^1^* | - | Testis | - |
| rs145982353 | - | - | - | *MIR527* | Micro RNA | - | - |
| rs6075980 | - | - | - | *ZCCHC3^1^* | DNA and RNA binding | - | - |
| rs148653405 | - | - | - | *GPCPD1^2^* | Glycerophospholipid catabolic process | - | - |
| 1. Intergenic variant, gene listed is nearest. | | | | | | | |
| 2. Intergenic variant, and in linkage disequilibrium (r2>0.8) with variant within gene. | | | | | | | |
